# Machine Learning Based Digital Assessment of Mild Cognitive Impairment Using Hand Movements during the Trail Making Test

**DOI:** 10.64898/2026.01.05.26343443

**Authors:** Gustavo E. Juantorena, Gianluca Capelo, Betsabe D. Leon Vallejos, Agustin Ibáñez, Agustin Petroni, Waleska Berrios, María Cecilia Fernández, Juan E. Kamienkowski

## Abstract

One of the objectives of digital neuropsychology is to apply computational methods to improve the accuracy of traditional assessments. We created and evaluated a computerized TMT (cTMT) that preserves its original structure and records high-resolution mouse trajectories. Seventy-four older adults (41 with mild cognitive impairment and 33 healthy controls) completed the cTMT and a standard diagnostic battery. We also developed NeuroTask, a Python library that extracts features from cursor time series, including reaction times, speed and acceleration metrics, trajectory deviations, and state-based measures. We compared demographic, digital, and non-digital models using nested cross-validation and non-parametric permutation tests. Demographic models provided only modest discrimination (AUC = 0.56). Digital hand features improved performance (AUC = 0.65), and combining them with demographics reached an AUC of 0.70, which approached the performance of the neuropsychological battery used to define the diagnosis (AUC = 0.76). In complementary regression analyses with digital plus demographic features, we obtained significant predictions for five of seven target scores: MMSE, Digit Symbol, TMT-A, TMT-B, and Forward Digit Span. These results indicate that fine-grained hand-movement features from the cTMT provide useful information for classifying mild cognitive impairment and for predicting multiple neuropsychological scores.

## Introduction

The World Health Organization (2017) projects that the global burden of dementia will nearly triple within the next thirty years. This neurological syndrome encompasses diverse disorders that progressively impair memory, cognition, and daily functioning, with considerable heterogeneity in underlying causes and clinical manifestations (Mukadam et al., 2024). However, these figures mask important regional disparities, with low- and middle-income countries, particularly those in Latin America, having the highest dementia prevalence rates and the sharpest increases, creating significant challenges in regions where healthcare resources are scarce (Aranda et al., 2021; Baez et al., 2023; Custodio et al., 2017; Kalaria et al., 2024). Alzheimer’s disease (AD), accounting for approximately 70% of dementia cases, is now defined as a biological continuum where neuropathological changes precede clinical symptoms by years (Jack Jr. et al., 2024). This framework emphasizes the importance of early detection. In this context, Mild Cognitive Impairment (MCI), previously considered merely prodromal, is now recognized as part of this AD continuum. Though not all individuals with MCI progress to dementia, risk remains high: approximately 80% advance within six years (Busse et al., 2006; Petersen & Negash, 2008).

In recent decades, advancements in computational power and the increased availability of digital devices have enabled clinicians and researchers to explore new applications of these tools in neuropsychology and neurology. These developments have given rise to the fields of digital neuropsychology (Germine et al., 2019) and computational psychiatry (Montague et al., 2012). In this context, it is important not only to adapt classic paper-and-pencil tests but also to consider, on the one hand, the most robust ways to administer them to obtain more information in less time with fewer resources and, on the other hand, the most efficient tools to extract relevant information for clinical purposes.

The Trail Making Test is one of the most used neuropsychological screening tests, which has been used worldwide for clinical assessment and research for over eighty years (Bowie & Harvey, 2006; Lange et al., 2005; Rabin et al., 2007; Reitan, 1958; Salthouse, 2011; Soukup et al., 1998). There are two parts. In Part A, the participant must connect the numbers in ascending order from 1 to 25 on a paper sheet. In part B, the circles contain a combination of letters (A to L) and numbers (1 to 13). The numbers and letters are placed in a semi-random order to avoid overlapping lines drawn by the participant (Linari et al., 2022). The primary variables of interest are the total times for Parts A and B. It is sensitive to executive function (EF) impairments in multiple clinical populations (Ashendorf et al., 2008; Giovagnoli et al., 1996; Periáñez et al., 2007). Different executive processes are thought to be associated with performance in the TMT, including inhibitory control, working memory, and attention (Arbuthnott & Frank, 2000; Kortte et al., 2002; Sánchez-Cubillo et al., 2009).

Early efforts to digitize the Trail Making Test (TMT) laid the groundwork for leveraging computational methods in cognitive assessment. Dahmen and collaborators (2017) introduced a digital version of the TMT and applied machine learning techniques to evaluate its predictive validity across clinical and cognitive outcomes. Recent developments in digital adaptations have shown promise in enhancing cognitive assessment across various populations and modalities. Park and Schott (2022) compared traditional paper-based and computerized TMTs in young and older adults, finding that digital versions can sensitively detect age-related differences. Several studies have employed digital tools for early detection of neurodegenerative conditions: Kobayashi et al. (2022) used drawing-based digital tasks, including TMT, to detect Alzheimer’s disease, while Chudzik et al. (2024) analyzed online TMT dynamics to differentiate patterns in Parkinson’s disease. Du et al. (2022) applied hidden Markov models to digital TMT sequences, extracting cognitive markers beyond total time. Verma and Dhyani (2025) validated an Android-based TMT app for both healthy and depressed individuals, supporting mobile assessments. Building on this trend, Lara-Garduno and collaborators (2022) introduced *SmartStrokes*, a tablet-based TMT interface. Their system applies line-segment-level classification models to detect MCI-related sketching behavior, and research led by Del Gatto (2025) reported a set-size effect in a novel digital TMT, providing insights into the mechanisms of visual search.

Building on these advances, recent research has increasingly leveraged machine learning (ML) to analyze multimodal behavioral data, including eye movements, motor behavior, and digital interactions, for the early detection of mild cognitive impairment (MCI) and Alzheimer’s disease (AD). For example, Revankar et al. (2025) demonstrated that smartphone-based behavioral profiling, incorporating fine-grained motor and interaction data, can distinguish between AD and Lewy body dementia in real-world conditions. These results are consistent with broader efforts in neuroscience and psychiatry that apply artificial intelligence to extract latent cognitive markers from naturalistic and high-dimensional data, as discussed in recent studies (Badrulhisham et al., 2024; Chekroud et al., 2021). Using digital markers based on hand and eye movement, Kim and co-authors (2023) trained Machine Learning models for early screening of mild cognitive impairment, showing the advantages of using new technologies. Additionally, research led by Dörr (2025) assessed a semantic verbal fluency task in participants with mild cognitive impairment or healthy aging. Then, they extracted speech features and trained a machine learning classifier on them to examine practice effects.

This shift from static test scores to dynamic, interpretable signals suggests that machine learning applied to ecologically valid modalities such as eye tracking, voice, and hand behavior may significantly improve early diagnosis and individualized monitoring of cognitive decline.

Our primary goal is to investigate whether fine-grained digital markers derived from the computerized Trail Making Test (cTMT) can support the early detection of mild cognitive impairment (MCI). We aim to capture subtle cognitive differences by combining classic completion time measures with detailed hand movement features, such as trajectory deviations, movement dynamics, and cursor kinematics. We evaluate the diagnostic value of these digital features by comparing their predictive performance with that of demographic information and standard neuropsychological tests. Additionally, we examine whether cTMT features can predict participants’ scores on non-digital neuropsychological tasks to assess the extent to which digital biomarkers can approximate established clinical evaluations. Ultimately, we demonstrate that digital markers obtained using widely accessible hardware, such as a standard computer and mouse, can complement or partially replace traditional assessments performed by health professionals, offering a scalable and low-cost tool for cognitive screening.

## Methods

### Participants

A total of 85 participants, 48 with Mild Cognitive Impairment (MCI) and 37 Healthy Controls (HC), were initially recruited, but after applying exclusion criteria, 74 participants (41 MCI) remained in the final analysis. Exclusions included five participants who reported difficulties using the computer mouse and were unable to connect at least four items during the practice trials, and six participants who did not complete at least one trial of each type (A or B).

All participants had normal or corrected-to-normal vision and performed the task using a computer mouse with their dominant hand. Individuals with prior diagnoses of neurological (e.g., Parkinson’s disease) or vascular conditions were excluded. Written informed consent was obtained from all participants, and the study protocol was approved by the Institutional Review Board of Hospital Italiano de Buenos Aires (ID: 11543) in agreement with the Helsinki declaration.

Patients were diagnosed with MCI based on an interview, an informant report, a neuropsychological evaluation that showed a performance of -1.5 SD in at least two tests in one cognitive domain (attention, memory, language, executive functions, or visuospatial skills), and those having activities of daily living preserved (Jak et al., 2009). A conservative criterion was used to determine the engagement of a cognitive domain, thus identifying the neuropsychological pattern as either uni- or multi-domain.

In addition to taking our digital test, participants completed a series of tasks that were used for their diagnostic evaluation. Those tasks were: (1) Mini-Mental State Examination (MMSE), (2) Trail Making Test (TMT)-A and -B, (3) Digit Symbol, (4) Forward and Backward Span, and (5) Clock Drawing Test.

No significant demographic differences were observed between the control (n = 33) and MCI (n = 41) groups (Table 1).

**Table 1.** Demographics. Fisher’s exact test indicated no association between group and sex. Independent samples t-tests revealed no significant differences in age or years of education between groups.

|  | HC | MCI | p-value |
| --- | --- | --- | --- |
| Age | 73 ± 8 | 76 ± 7 | 0.078 |
| Male / Female | 8 / 25 | 11 / 30 | 1.000 |
| Years of education | 14 ± 3 | 14 ± 4 | 0.683 |

### Computerized TMT (cTMT)

Building on our previous work (Linari et al., 2022), we adapted the Trail Making Test (TMT) (Bowie & Harvey, 2006) to a digital format using Python (v3.10.16; Python Software Foundation, 2024) and PsychoPy (*v2022.1.1*; Peirce et al., 2019), tailoring it to the characteristics of the target population. The task preserved the structure of the original TMT. In each trial, participants were required to connect 15 items in sequential order. In TMT-A, the items consisted of numbers from 1 to 15. In TMT-B, the items included both numbers (1 to 8) and capital letters (A to G), which participants had to connect in an alternating sequence, beginning with number 1 (i.e., 1, A, 2, B, etc.) (Fig. 1A). The complete task included 20 trials, alternating between TMT-A and TMT-B, starting with TMT-A. Each trial began when participants clicked a central fixation cross, after which the stimuli appeared. Trials lasted a maximum of 25 seconds and ended either when the participant selected the final item in the sequence or when the time limit was reached. Mouse movements were continuously recorded as x and y screen coordinates with associated timestamps (t), enabling reconstruction of the full trajectory (Fig. 1B).

**Figure 1.**
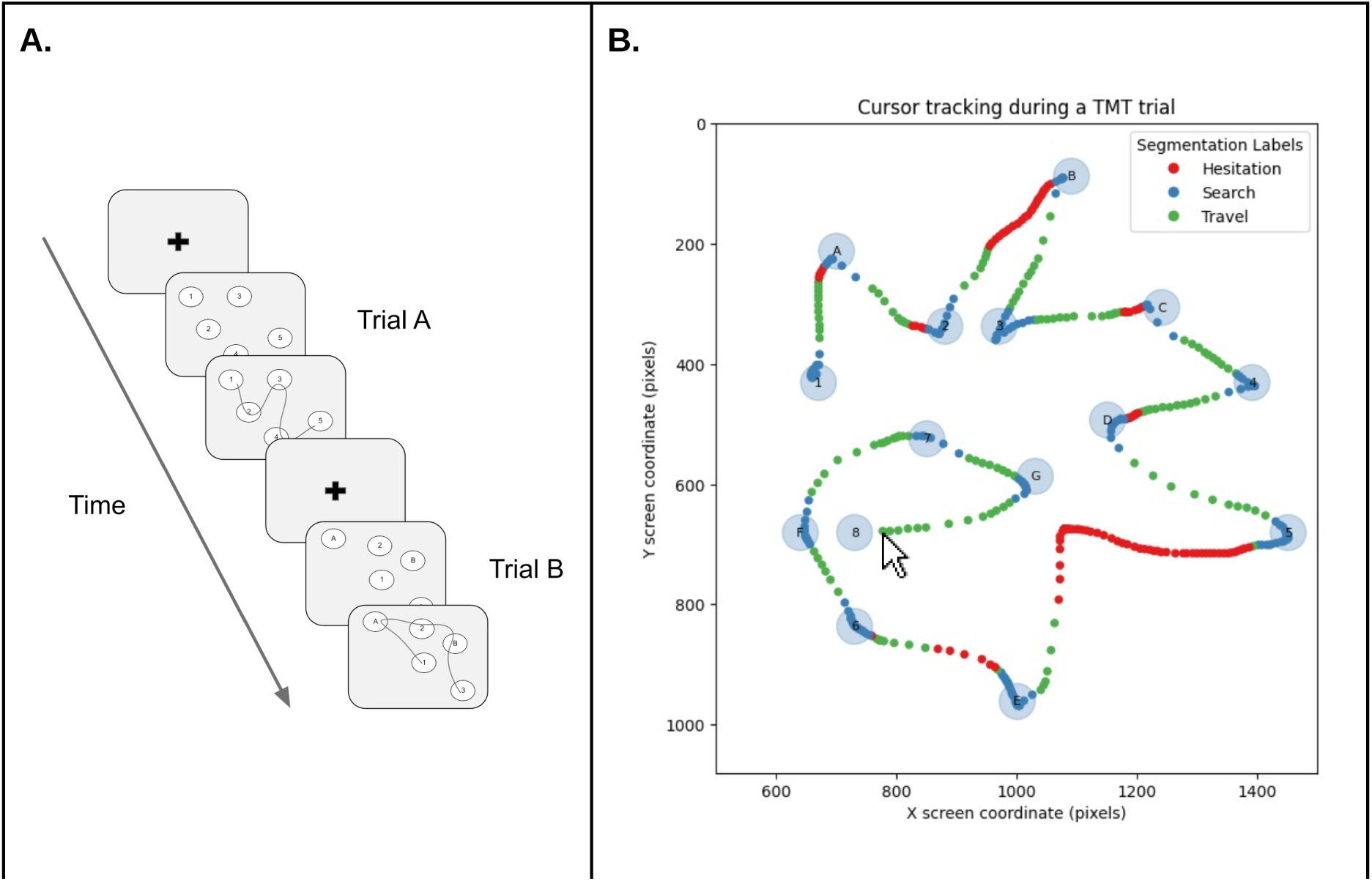
TMT Paradigm and mouse movement processing. **A.** Participants alternated between TMT-A and TMT-B trials, each beginning with a central fixation cross. In Part A, items were numbers 1-15 in ascending order; in Part B, items were numbers (1-8) and letters (A-G) arranged in alternating sequence. **B. Mouse movements during a TMT-A trial**. Participant’s cursor path during the cTMT. The numbered circles represent the targets, and the scatter plot shows the sequential path of the participant’s mouse between them. The color of the points indicates the state: hesitation (red), search (blue), or travel (green).

### Feature extraction from hand measurements

We extracted several hand measurements based on mouse movement time series using NeuroTask (https://github.com/NeuroLIAA/neurotask), a Python package that we created for this purpose. This tool allows us to extract multiple metrics from the cursor time series. The complete measures are presented in Table S1 of the supplementary materials.

First, we obtained classic TMT measures such as the total time in trial, the number of correctly and incorrectly touched targets, the number of crosses over previous strokes, among others. Next, we estimated the degree of deviation from the ideal path. We defined this as the trajectory resulting from connecting the centers of each target with straight lines in the correct order. From this benchmark, we derived two metrics: the cumulative difference between the actual and ideal trajectories (in terms of point-to-point distance), and the area between the two trajectories, as an integrated measure of spatial deviation. Additionally, we extracted kinematic metrics based on the cursor path, including average speed, maximum speed, and characteristics derived from acceleration (e.g. zigzag amplitude, defined as the mean time difference between consecutive number and letter transitions in part B). We also incorporated metrics used in the literature, such as dwell time/intra-target time (time spent on targets), and its complement, inter-target time (time spent away from targets) (Thornton & Horowitz, 2020).

Inspired by the work of Lara-Garduno and collaborators (2022), we segmented each trial into different states that aim to represent the subject’s actions (Fig. 1B). The *Search* state refers to periods when the participant is looking for the next target. This includes points within a target or immediately after leaving it. However, the subject’s speed cannot exceed a subject-specific speed threshold, which was calculated as the median cursor speed during the movement from the first to the second target. This condition avoids classifying points outside the target due to involuntary movement or proximity. The *Travel* state describes when the subject moves decisively in a given direction, including points that do not meet the search condition and exceed the speed threshold; *Hesitation* represents moments of doubt, trajectory correction, or temporary loss of the target, including segments immediately after a travel state in which the subject slows down below the threshold. For each state, we extracted specific metrics, such as distance traveled, elapsed time, and average speed, allowing for a more detailed characterization of the subject’s resolution.

Finally, we considered a trial valid even if the final target was not reached. To ensure comparability across participants, we set a threshold of eight targets. (more than fifty percent of the sequence) and use only the trajectory up to the last correctly reached target to calculate it. This yielded two sets of features: one restricted to the trajectory up to the last correctly reached target, and one without this limitation (variables labeled with the prefix “Complete” in Table S1).

### Machine learning pipeline

We implemented a nested cross-validation (CV) framework to obtain unbiased performance estimates across all models (Fig. 2). The outer loop used Leave-One-Out CV (LOO) to evaluate generalization performance. In each iteration, one subject was held out as the test fold, and the remaining subjects formed the training set. All preprocessing steps, including standardization and univariate feature selection, were performed exclusively on the training data and then applied to the test fold to prevent data leakage (Sasse et al., 2025). Feature selection was carried out using the SelectKBest method from scikit-learn (v1.7.1; Pedregosa et al., 2018), which ranks features according to their univariate statistical relationship with the target variable. The ANOVA F-test (f_classif) was used as the scoring function, and the top k features (k ≤ 20) were retained based on their F-scores. When fewer than 20 features were available, all were included in the model.

**Figure 2.**
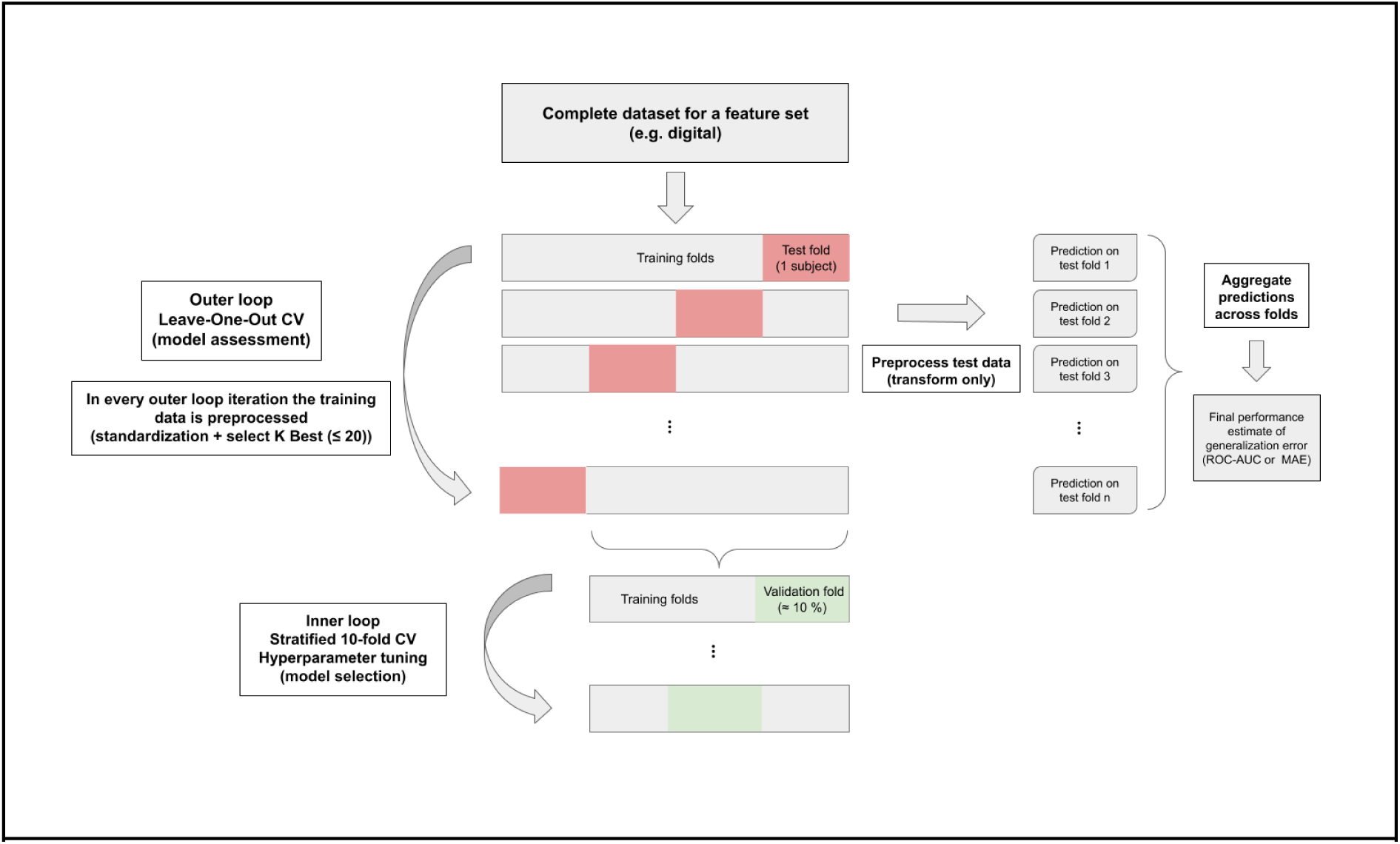
Machine Learning Pipeline. Representation of the nested cross-validation procedure used in this study. The outer loop (Leave-One-Out CV) was used to estimate model generalization performance. In each iteration, one subject was held out as the test fold, while the remaining subjects formed the training folds. Preprocessing, including standardization and univariate feature selection with SelectKBest (≤20 features), was performed exclusively on the training data and then applied to the test fold to prevent data leakage. Within each outer training set, stratified 10-fold cross-validation (inner loop) was performed for hyperparameter tuning and model selection. Predictions for the left-out subjects were aggregated across outer folds to compute the final performance estimates, using ROC-AUC for classification and MAE for regression.

Within each outer training set, we performed stratified 10-fold CV (inner loop) for hyperparameter tuning and model selection. Hyperparameters were optimized via grid search (GridSearchCV) using the area under the ROC curve (ROC-AUC) for classification and mean absolute error (MAE) for regression based on the recommendations of Poldrack and collaborators (2020). After optimal parameters were selected, the pipeline was retrained on the entire outer-training partition and evaluated on the left-out subject. Predictions were aggregated across folds to compute the final performance estimate. This methodology is crucial for obtaining unbiased performance estimates, as it strictly separates the data used for model selection from the data used for performance evaluation (Raschka, 2020).

The “no-free-lunch” theorems (Wolpert, 1996) assert that no single algorithm consistently outperforms all others across every problem domain. Accordingly, we evaluated several alternative models to determine the most suitable approach for our data (see Results). Complete hyperparameter grids for all models are provided in Table S2 of the Supplementary Materials.

### Feature importance: SHapley Additive exPlanations (SHAP) approach

We utilized the SHapley Additive exPlanations (SHAP) approach to get a deeper understanding of the models’ behavior (Lundberg & Lee, 2017). We then computed the SHapley values within each outer LOOCV fold. For each fold, the chosen model with its optimized hyperparameters was refitted on the training set. Then, shapley values were calculated for the held-out subject, using the training set as background. In classification tasks, explanations were obtained from the predicted probability of the positive class and in regression tasks, from the raw prediction. Since feature selection (SelectKBest) could lead to different subsets of features being included in different folds, we aggregated the values by averaging the absolute contribution of each feature across all folds in which it appeared. This procedure yielded overall importance scores that reflect both the magnitude of a feature’s contribution and its stability across folds.

### Demographic baseline models

To establish a lower-bound reference, we first trained models using only three demographic variables: sex, age, and years of education (see Table 1). These variables are commonly associated with cognitive performance and are usually included in normative corrections for neuropsychological test scores. While they are not specific markers of cognitive impairment, they provide a useful benchmark for evaluating the added value of task-based digital features.

### Neuropsychological benchmark models

To establish an upper-bound reference, we trained models using the same neuropsychological assessments that were used for the original clinical diagnosis of MCI, as described in the “Participants” section. These measures included the Mini-Mental State Examination (MMSE), Trail Making Tests A and B, the Digit Symbol Substitution Test, Forward and Backward Digit Span, and the Clock Drawing Test. During the clinical evaluation, these standardized tests were supplemented with an unquantified clinical interview and informant report. We expected this model to achieve the highest classification performance, which would serve as a ceiling against which the contribution of digital biomarkers could be compared.

### cTMT digital features models

Finally, we used hand-derived features from the computerized Trail Making Test (cTMT) to train machine learning models that capture fine-grained motor behavior associated with cognitive decline in participants with mild cognitive impairment (MCI) compared with healthy controls. We extracted these features using NeuroTask (https://github.com/NeuroLIAA/neurotask). The library processes mouse movement time series to compute metrics that characterize task performance and movement dynamics.

The extracted variables included classic TMT measures, such as total completion time, number of correct and incorrect touches, and trajectory crossings, as well as spatial deviation metrics, including cumulative distance and area between the actual and ideal paths. In addition, we computed kinematic measures (average and peak speed, and acceleration-based indices) and state-segmented features derived from three behavioral states: Search, Travel, and Hesitation, identified within each trial (Fig. 1B). For each state, we calculated specific measures of distance traveled, elapsed time, and mean speed, allowing a detailed characterization of the behavior during the task. A complete list of extracted features is provided in Supplementary Table S1.

## Results

### Exploratory Analysis

We divided the data for each subject into three feature sets: demographics (sex, age, and years of education); non-Digital (six features from standard tests used in the diagnostic pipeline, such as TMT A and B and MMSE); and digital (100 features extracted from our cTMT, such as time to complete, number of correct trials, and distance from the ideal path).

As described in the Methods section, the MCI and HC groups did not differ significantly in age, sex, or years of education. In contrast, significant group differences emerged in the non-digital tests performed by neuropsychologists for diagnosis. Significant differences in performance on several variables were observed, assessed by independent-samples t-tests. MMSE scores were lower in the MCI group (t = 4.26, p < 0.001), consistent with global cognitive decline. Similarly, MCI participants had significantly longer TMT-A (t = −3.98, p < 0.001) and TMT-B (t = −5.37, p < 0.001) completion times, reflecting reduced processing speed and executive functioning. Additional differences were observed on the Digit Symbol Test (t = 2.99, p = 0.004), the Backward Digit Span Test (t = 2.61, p = 0.011), and the Clock Drawing Test (t = 2.50, p = 0.015), indicating poorer performance in the MCI group. These results confirm that the two groups differed consistently across the reference battery, providing a valid benchmark for evaluating the digital measures in our study.

Features extracted from the cTMT also revealed several significant group differences. In Part A, MCI participants showed larger deviations from the ideal path (t = −2.12, p = 0.038), longer hesitation times (t = −2.05, p = 0.045), and longer intra-target times (t = −2.35, p = 0.022). In Part B, participants with MCI exhibited more complete correct touches (t = 3.36, p = 0.001), longer complete time in trial (t = –2.21, p = 0.03), and higher complete correct touches (t = 2.23, p = 0.029). Additionally, the total number of correct B-type trials was significantly higher in the HC group (t = 2.95, p = 0.004). These exploratory results suggest that, beyond the raw completion time, fine-grained digital markers from the cTMT can capture subtle differences between groups.

### Diagnostic group classification

We trained four machine learning algorithms to predict the diagnostic group of the participants: Random Forest (RF), Support Vector Machine (SVM), Logistic Regression (LR), and XGBoost (XGB). Each algorithm was trained and evaluated using five feature sets: demographic variables, digital features derived from the cTMT, non-digital neuropsychological measures, and two combined sets (demographic plus digital and demographic plus non-digital).

For each feature set, we selected the model with optimal hyperparameters, as determined by the nested cross-validation procedure. The corresponding ROC curves were plotted (Fig. 3A). The AUC values summarize the overall discriminative ability and highlight the relative contribution of each feature set.

**Figure 3.**
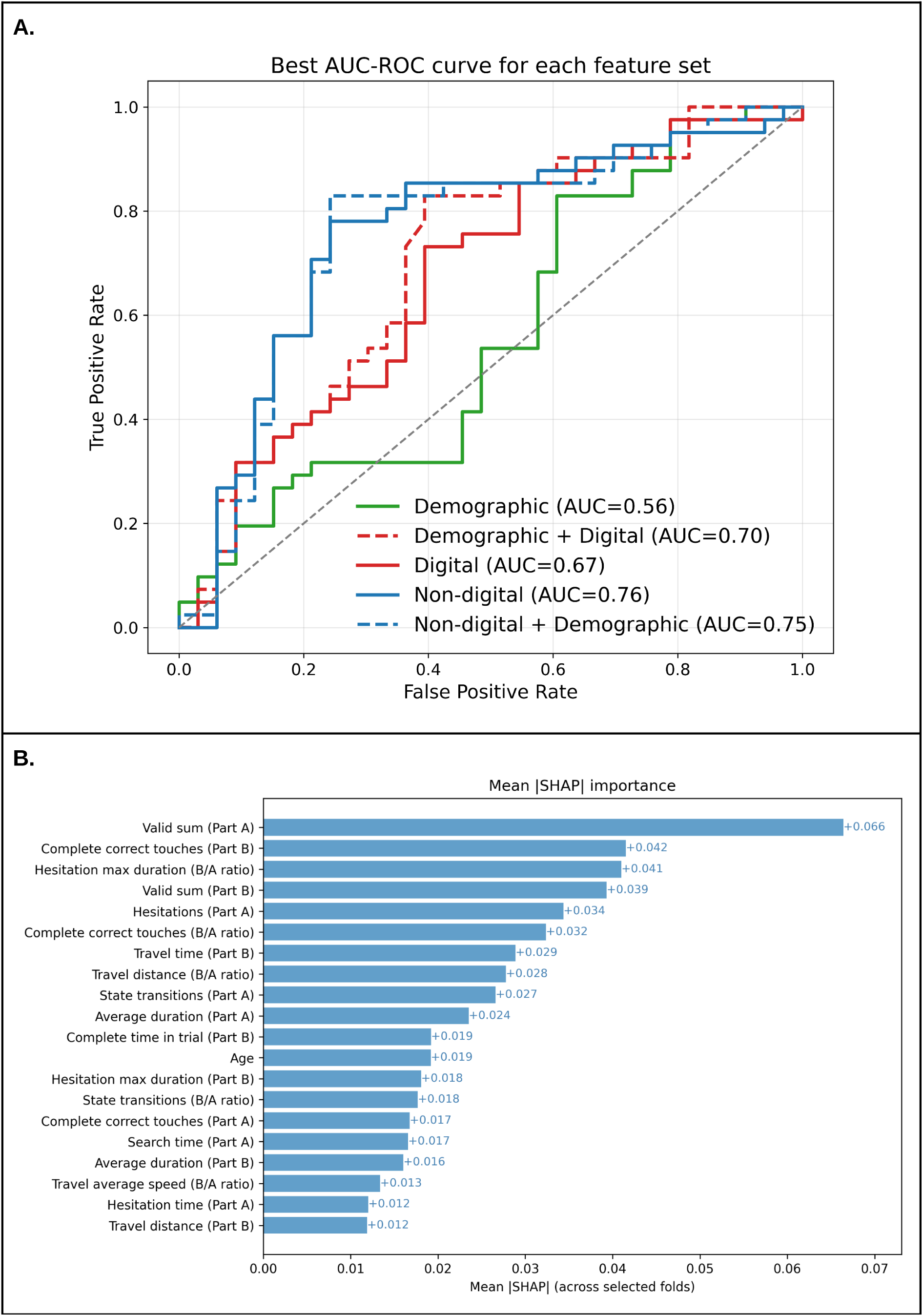
Diagnostic group classification performance and feature importance. **A.** Receiver operating characteristic (ROC) curves comparing classification performance across feature sets in the full sample (N = 74, 41 MCI). Area under the curve (AUC) values are reported for the bestperforming model within each feature set. Demographics alone provided only modest discrimination (RF, AUC = 0.56, p = 0.189). Digital features from the computerized TMT improved performance (SVM, AUC = 0.67, p = 0.007), and combining demographics with digital features further increased accuracy (SVM, AUC = 0.70, p = 0.002). Non-digital neuropsychological measures yielded the highest discriminative value (RF, AUC = 0.76, p = 0.001), with demographics adding no further benefit (RF, AUC = 0.75, p = 0.001). **B.** Mean absolute SHAP values (mean |SHAP|) for the classification model. Importance scores were averaged across folds, accounting for varying feature subsets due to feature selection.

To assess whether model performance exceeded chance level, we performed non-parametric permutation tests with 1,000 label shuffles for each feature set and classifier. The empirical p-value was defined as the proportion of permutations yielding an AUC greater than or equal to the observed one. As expected, demographic variables alone provided only modest discrimination (RF, AUC = 0.56, p = 0.189), indicating that performance was not significantly better than random guessing. In contrast, digital features derived from the computerized TMT showed a clear improvement (SVM, AUC = 0.67, p = 0.007). Combining demographics with digital features further increased performance (SVM, AUC = 0.70, p = 0.002), highlighting the added value of the cTMT beyond demographic information alone. As a benchmark, non-digital neuropsychological measures achieved the highest overall discriminative performance (RF, AUC = 0.76, p = 0.001). Adding demographics to these measures almost did not change the AUC (RF, AUC = 0.75, p = 0.001).

Collectively, these results support the interpretation that digital features, when integrated with demographic data, enhance classification performance beyond the demographic information, yielding discrimination levels comparable to those obtained with traditional neuropsychological assessments.

Global feature importance was expressed as the mean absolute SHapley value across folds (Fig. 3B). In brief, Shapley values were computed for each held-out subject using the refitted model from that fold’s training set, then aggregated across folds by averaging the absolute contributions of each feature where it appeared after feature selection. These values quantify each feature’s contribution to the model’s predictions, where higher absolute values indicate greater influence on the predicted probability of the positive class (MCI). Age, from the demographic feature set, also appears in the twelfth position. The most influential predictors, as indicated by their high SHAP values, were the number of trials with at least eight targets correctly connected (*Valid sum*) in Part A, the number of correct target touches without a target threshold (*Complete correct touches*) and *Valid sum* in Part B, and the ratio of the longest hesitation durations (*Hesitation max duration ratio*) in both Parts B and A. Another top predictor was *state transitions*, which quantify how often the cursor switches between the hesitation, search, and travel states.

### Neuropsychological test score prediction

Consistent with the *Diagnostic Group Classification* section, seven machine learning algorithms were trained to predict the scores from the non-digital neuropsychological tasks: Linear Regression (LR), Ridge Regression (RR), Lasso (Lasso), Elastic Net (EN), Support Vector Regression (SVR), Random Forest Regression (RFR), and XGBoost Regression (XGBR). Each algorithm was trained using digital features in combination with demographic data to predict a single non-digital neuropsychological task at a time. We followed the same pipeline as in the classification setting. This included nested cross-validation, hyperparameter optimization through grid search (Fig. 2), and a permutation procedure to assess significance. The complete set of parameters explored in the grid search is provided in Table S3 of the Supplementary Materials.

We obtained significant predictions for five out of the seven non-digital test scores evaluated (Table 2). For the MMSE, the best model was RFR (MAE = 1.45, non-parametric permutation test, p = 0.004). The Digit Symbol test was best predicted by Lasso (MAE = 9.47, p = 0.001). TMT-A was predicted with EN (MAE = 11.95, p = 0.001), and TMT-B with SVR (MAE = 43.09, p = 0.001). Forward digit span was predicted with Elastic Net (MAE = 0.83, p = 0.027). Conversely, the Backward Digit Span and Clock Drawing Test did not reach significance, the best model was SVR in both cases (MAE = 1.13, p = 0.419, and MAE = 0.73, p = 0.837, respectively).

**Table 2:** Prediction of individual (Target) tests in the demographic + digital feature set. RFR: Random Forest Regressor, SVR: Support Vector Regressor, SD and IQR: Standard Deviation and Interquartile Range of Target values, MAE: Mean Absolute Error of predicted vs target values, p-value estimated from permutation test (see Methods).

| Target | Best model | Target values |  | Predicted values |  |
| --- | --- | --- | --- | --- | --- |
|  |  | SD | IQR | MAE | p-value |
| MMSE | RFR | 1.9 | 2.0 | 1.45 | 0.004 |
| Digit Symbol | Lasso | 13.7 | 16.8 | 9.47 | 0.001 |
| TMT-A | EN | 17.2 | 20.1 | 11.95 | 0.001 |
| TMT-B | SVR | 64.0 | 69.8 | 43.09 | 0.001 |
| Forward Digit Span | EN | 1.0 | 1.0 | 0.85 | 0.027 |
| Backward Digit Span | SVR | 0.8 | 1.8 | 0.62 | 0.419 |
| Clock Drawing | SVR | 0.5 | 0.0 | 0.29 | 0.837 |

We also calculated the standard deviation (SD) and interquartile range (IQR) of the target values to provide a reference for interpreting the mean absolute error (MAE) in relation to the variability inherent to each neuropsychological measure. This allows us to contextualize the prediction errors against the real spread of the scores. In every case, the MAE is smaller than the SD, and even smaller than the IQR. It would be noticed that the clock drawing test suffers from a lack of variability between quartiles one and three, resulting in an IQR of zero, which is the product of the variability of the score being very small, with 76% of the participants receiving a score of 3 (on a scale of 1-3). Taken together, these results suggest that our models can capture informative patterns across most neuropsychological measures. Our models are particularly accurate at predicting scores on tests with sufficient variability.

Similar to the *Diagnostic Group Classification* analysis, we studied the global feature importance by observing the mean absolute SHapley value across folds (Lundberg & Lee, 2017) in the significant cases (Fig. 4). In all these cases Age appears within the top twelve positions, and in MMSE, Digital symbol, and TMT-A, it appears within the top three. For the Digit Symbol task, hesitation time and mean negative acceleration were found to be the dominant factors (Fig. 4A). This pattern is consistent with the task’s dependence on working memory, as longer pauses may reflect moments when participants search the screen due to difficulty recalling the next item (Linari et al., 2022; Wölwer et al., 2003). In TMT-A, age was the strongest predictor, followed by the number of correct touches in part A, which aligns with findings from both the traditional and digital versions (Fig. 4B). Performance and speed features also contributed, with hesitation-related metrics showing moderate influence. In TMT-B, the strongest features were derived from part B, consistent with findings from the paper-based version (Fig. 4C). Accuracy and timing measures dominated, and distance from ideal also appeared among the top features. In the Forward Digit Span, the leading features were mainly related to movement speed and timing, with both parts contributing similarly (Fig. 4D). It is worth noting that two features related with zigzag behavior amplitude were also present. Finally, for the MMSE, feature importance exhibited a more homogeneous distribution across domains, similar to the distribution of features for the MCI versus HC classification (Fig. 3B and 4E). Although age and the ratio of state transitions between parts B and A showed the highest SHAP values, several other variables contributed meaningfully, including speed-related, performance-based, and state-derived measures such as hesitation time and total hesitations (Fig. 4E). This pattern suggests that MMSE performance is influenced by a broader set of motor and behavioral features, rather than being driven by a single dominant construct. This is consistent with a screening battery which assesses various domains, such as orientation, attention, memory, and language (Tombaugh & McIntyre, 1992).

**Figure 4.**
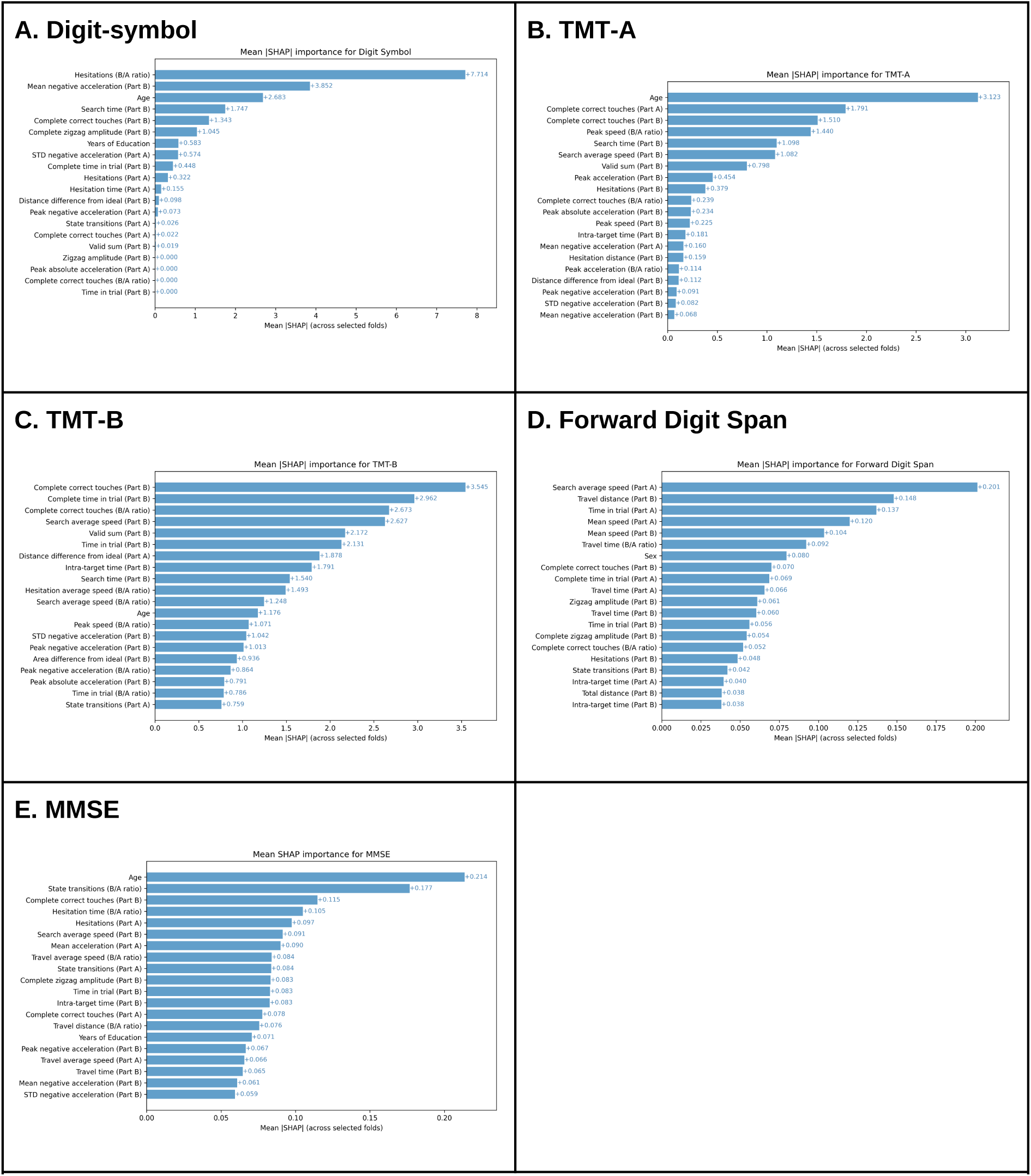
Mean absolute SHAP values (mean |SHAP|) for the significant regression models. (see Table 2). **A.** Digit Symbol. **B**, TMT-A, **C.** TMT-B, **D.** Forward Digit Span, and **E.** MMSE.. All models used the feature set that combines Digital and Demographic features. Importance scores were averaged across folds, accounting for varying feature subsets due to feature selection.

## Discussion

This study evaluated whether fine-grained hand movement features extracted from a computerized Trail Making Test (cTMT) could discriminate between individuals with mild cognitive impairment and cognitively healthy controls. Our approach incorporated three complementary sources of information: (i) demographic variables (age, sex, years of education), (ii) standardized neuropsychological assessments used in the original MCI diagnosis, and (iii) High-resolution digital features derived from mouse-based hand trajectories. By systematically comparing models using these three feature sets within a leave-one-out nested cross-validation machine learning pipeline, we were able to quantify the potential added diagnostic value of digital motor biomarkers relative to both minimal (demographics only) and maximal (full neuropsychological battery) reference points.

Additionally, we used the same pipeline to predict continuous neuropsychological scores based on cTMT features. This analysis revealed that the fine-grained motor patterns captured during test performance contain information useful not only for classification, but also for identifying systematic associations with individual variability across different cognitive domains. Using SHAP analysis, we found that age ranked among the top three predictors in the MMSE, Digit Symbol, and TMT-A models, in line with prior reports linking age to broad neuropsychological performance (Hamdan & Hamdan, 2009; Hoyer et al., 2004)

More importantly, distinct yet interpretable feature patterns emerged for each test. For the Digit Symbol task, hesitation time and mean negative acceleration were the most influential features, consistent with their links to working memory and search-related pauses. The MMSE displayed a more evenly distributed profile, including state transitions and performance-based variables, which aligns with its multidomain structure. For TMT-A and TMT-B, predictive features derived mainly from their corresponding parts, with accuracy, completion time, and trajectory efficiency emerging as key contributors. Finally, in the Forward Digit Span, speed and timing features were again prominent, while zigzag amplitude appeared among the predictors, suggesting that temporal alternation between targets may reflect attention and short-term memory maintenance. Together, these results support the view that digital biomarkers derived from hand movement trajectories can serve as sensitive indicators of cognitive performance.

Given the simplicity of the required hardware, this approach could also be implemented remotely, enabling broader accessibility and scalability. Moreover, these findings complement other work exploring multimodal approaches to digital TMTs. For example, eye-tracking methods have been integrated into similar frameworks. In previous research, we found that combining hand- and eye-tracking metrics during a computerized TMT correlated with specific executive function components (Linari et al., 2022). Similarly, the ETMT tool (Chandrasekharan et al., 2023) employs eye tracking and adaptive neuro-fuzzy inference to produce an overall cognitive-impairment score, while Jyotsna et al. (2020) used gaze-tracking data to classify participants into low, medium, or high impairment groups. Collectively, these approaches illustrate how the TMT can be adapted into digital and multimodal contexts that capture cognitive processes in more detail

Compared with previous studies that used digital TMT movement-based features for MCI classification, our work differs in its scope, methodology, and analytical approach. For instance, Hanczár et al. (2022) analyzed short mouse movement segments using proprietary software developed by the company with which the authors are affiliated and a decision tree model trained with synthetic data augmentation and validated through repeated random splits. In contrast, our analysis relies on complete trial trajectories and standardized feature extraction through our open-source NeuroTask library. We also implemented a more rigorous evaluation using nested cross-validation and permutation testing. Lara-Garduno et al. (2022) developed SmartStrokes, a tablet-based system that incorporates sketch recognition. The authors also used regression models to predict neuropsychological test scores, reporting an average MAE of approximately 2.4 (sd. 0.110) in MoCA, whereas our MMSE predictions achieved a lower value of 1.45 (p = 0.004). Our analysis therefore demonstrates comparable or improved accuracy. Together, these findings suggest that both stylus- and mouse-based approaches can capture meaningful cognitive variability, while our framework emphasizes transparency, openness, and methodological rigor.

The rapid proliferation of machine learning models for MCI diagnosis and progression prediction has often outpaced methodological rigor. Reviews have highlighted that many studies omit critical details about feature selection, validation procedures, or reproducibility, limiting clinical translation (Pellegrini et al., 2018; Singh et al., 2024). By adhering to established reporting and validation standards (Poldrack et al., 2020; Sasse et al., 2025), the present work contributes to ongoing efforts to improve transparency and reliability in digital neuropsychology.

Importantly, the use of nested cross-validation maximized data use while maintaining an unbiased estimate of model performance. This approach is particularly suitable for small datasets, where holding out a separate test set would further reduce sample size and increase the variance of performance estimates (Varoquaux, 2018). Our goal was not to produce a deployable diagnostic model but to rigorously compare feature sets under consistent validation conditions.

Despite these contributions, several limitations should be acknowledged. First, performance in digital tasks may vary depending on the input device and its sensitivity, which can introduce unwanted variability. Although all participants in this study used the same mouse and display, this remains an important consideration for future implementations or cross-study comparisons. Second, the present work focused on a unimodal approach; combining hand movement with other modalities such as eye-tracking, speech, or physiological measures could provide a more comprehensive view of cognitive function. Third, while our sample size was typical for studies of this kind, it limits statistical power and generalizability, underscoring the need for replication in larger and more diverse cohorts.

A key strength of this approach lies in its low cost and compatibility with standard computing hardware, facilitating potential use in both clinical and remote settings. Unlike some standardized tests (e.g., MMSE), the cTMT can be freely implemented without licensing restrictions. Furthermore, the open-source release of NeuroTask library promotes methodological transparency and reproducibility, enabling other groups to extend or adapt the framework.

Mild cognitive impairment is a heterogeneous condition that remains challenging to define, identify, and monitor in clinical practice (Sabbagh et al., 2020). The growing adoption of digital neuropsychological tools, including computer-based versions of the Trail Making Test, reflects a broader shift toward data-rich and accessible cognitive assessments. However, the clinical utility of these approaches depends on balancing usability, interpretability, and rigorous validation.

Our results demonstrate that fine-grained hand movement trajectories obtained from a simple mouse-based task can meaningfully predict cognitive performance. By integrating open software, standardized pipelines, and transparent validation, this work contributes a replicable framework for the development of accessible, low-cost cognitive screening tools. Such approaches are particularly relevant for low-resource settings, common in Latin America, where affordable hardware and open methodologies can facilitate early and scalable cognitive assessment.

## Supporting information

SuppMat

## Data availability

The data used for the analysis are available at Open Science Foundation (OSF) repository: https://osf.io/nm9xy/overview?view_only=ed7edfe613d7488892362a1b3883a589

## Acknowledgments

We thank Mailen Guerra and Alejandro Masin for their contributions during the data collection.

## Author contributions

G.E.J., A.P., W.B., M.C.F., and J.E.K. designed the study; G.E.J. and B.S.V collected the data; W.B. and M.C.F. supervised the definition of MCI cases; G.E.J., G.C., and J.E.K. analyzed the data; G.E.J, G.C., and J.E.K. wrote the first version of the manuscript; and all authors revised the final version of the manuscript.

## Funding

The data collection was funded by the Seed Grant for Brain Health Leaders BL-SRGP2021-02 (BrainLat, Chile) and PICT 2018-02699 (Agencia I+D+i, Argentina).

## Code availability

All code used for the analyses is available at https://github.com/NeuroLIAA/tmt-analysis and the Neurotask repository https://github.com/NeuroLIAA/neurotask.

## Additional Information

The authors declare no competing interests.

## Notes

### Competing Interest Statement

The authors have declared no competing interest.

### Author Declarations

Written informed consent was obtained from all participants, and the study protocol was approved by the Institutional Review Board of Hospital Italiano de Buenos Aires (ID: 11543) in agreementwith the Helsinki declaration.

