## Supplementary material for "Machine Learning Based Digital Assessment of Mild Cognitive Impairment Using Hand Movements during the Trail Making Test": SuppMat

**Table S1: Hand features extracted from the mouse movement time-series. The same variables were computed separately for Part A, Part B, and for the B/A ratio, resulting in three variants of each feature.**

| Feature | Description |
|---|---|
| **Valid sum** | Indicates the number of valid trials (i.e., those with at least eight targets correctly connected). |
| **Total distance** | Total distance traveled by the cursor. |
| **Time in trial** | Total duration of the trial. |
| **Complete time in trial** | Total duration of the trial without threshold (i.e. 8 targets). |
| **Correct touches** | Number of correct target touches |
| **Complete correct touches** | Number of correct target touches without threshold (i.e. 8 targets). |
| **Mean speed** | Average cursor speed. |
| **SD speed** | Standard deviation of the cursor speed. |
| **Peak speed** | Maximum cursor speed observed. |
| **Mean acceleration** | Average acceleration of the cursor. |
| **SD acceleration** | Standard deviation of the cursor acceleration. |
| **Peak acceleration** | Maximum acceleration observed. |
| **Mean absolute acceleration** | Average of the absolute acceleration values. |
| **SD absolute acceleration** | Standard deviation of the absolute acceleration values. |
| **Peak absolute acceleration** | Maximum absolute acceleration observed. |
| **Mean negative acceleration** | Average of the negative acceleration values (deceleration). |
| **SD negative acceleration** | Standard deviation of the negative acceleration values. |
| **Peak negative acceleration** | Most negative acceleration value observed. |
| **Hesitation average speed** | Average hesitation speed. |
| **Hesitation distance** | Total distance traveled by the cursor in hesitation |
| **Hesitation time** | Total time spent in the hesitation state during the trial. |
| **Hesitation max duration** | Longest hesitation duration during the trial |

| | |
|---|---|
| **Travel distance** | Total distance traveled during travel segments. |
| **Travel time** | Total time spent during travel segments. |
| **Travel average speed** | Average cursor speed during travel segments. |
| **Search distance** | Total distance traveled during search movements. |
| **Search time** | Total time spent in the search state. |
| **Search average speed** | Average cursor speed during search movements. |
| **Inter-target time** | Sum of time elapsed between successive target touches. |
| **Intra-target time** | Sum of time spent within the area of a target before leaving it. |
| **Area difference from ideal** | Difference between the area of the actual path and the ideal path. |
| **Distance difference from ideal** | Difference between the traveled path length and the length of the ideal straight line connecting to the next target. |
| **State transitions** | Total number of transitions for one state (hesitation, search or travel) to another |
| **Complete zigzag amplitude** | Mean time difference between consecutive number and letter transitions in part B. |

**Table S2: Classification hyperparameters**

| Name | Hyperparameter | Values |
|---|---|---|
| **Random Forest Classifier** | n_estimators | 100, 200, 500 |
| | max_depth | None, 8, 16 |
| | min_samples_leaf | 1, 2, 5, 10 |
| | max_features | sqrt, log2 |
| **Support Vector Machine (SVC)** | kernel | linear, rbf |
| | C | 0.1, 1, 10 |
| | gamma (for rbf kernel) | "scale", "auto" |

| | | |
|---|---|---|
| **Logistic Regression** | penalty | l1,l2,elasticnet |
| | C | 0.1, 1, 10 |
| | solver | lbfgs, liblinear, saga |
| | l1_ratio (for elasticnet penalty) | 0.5 |
| **XGBoost Classifier** | n_estimators | 100, 200 |
| | max_depth | 3, 6 |
| | learning_rate | 0.1, 0.3 |
| | subsample | 0.8, 1.0 |
| | colsample_bytree | 0.8, 1.0 |

**Table S3: Regression hyperparameters**

| Name | Hyperparameter | Values |
|---|---|---|
| **Random Forest Regressor** | n_estimators | 100, 200, 500 |
| | max_depth | None, 8, 16 |
| | min_samples_leaf | 2, 5, 10 |
| | max_features | sqrt, log2 |
| **Support Vector Machine (SVR)** | C | 0.01, 0.1, 1, 10, 100, 1000 |
| | epsilon | 0.01, 0.1, 0.5, 1.0 |
| | gamma (for rbf kernel) | scale, auto |
| **Linear Regression** | No tunable hyperparameters | - |
| **Ridge Regression** | alpha | 0.0001, 0.001, 0.01, 0.1, 1.0, 5, 10.0, 100.0, 1000.0, 10000.0 |
| **Lasso Regression** | alpha | 0.0001, 0.001, 0.01, 0.1, 1.0, 5, 10.0 |
| **XGBoost Regressor** | n_estimators | 100, 200 |
| **Elastic Net** | alpha | 0.0001, 0.001, 0.01, 0.1, 1, 10, 100 |
| | l1_ratio | 0.05, 0.2, 0.5, 0.8, 0.95 |
